# Chest CT findings and outcomes of COVID-19 in second wave: A cross-sectional study in a tertiary care centre in Northern India

**DOI:** 10.1101/2023.03.17.23287423

**Authors:** Taranjeet Cheema, Amit Saroha, Arjun Kumar, Prasan Kumar Panda, Sudhir Saxena

**Affiliations:** Department of Internal Medicine (ID Division); Department of Radiology, AIIMS Rishikesh

**Keywords:** Corona Virus Disease 2019, Computed tomography score, Mortality, Pneumomediastinum, Pneumothorax

## Abstract

**Introduction:** The COVID-19 pandemic has posed a serious threat to global health, with developing nations like India being amongst the worst affected. Chest CT scans play a pivotal role in the diagnosis and evaluation of COVID-19, and certain CT features may aid in predicting the prognosis of COVID-19 illness.

**Methods:** This was a single-centre, hospital-based, cross-sectional study conducted at a tertiary care centre in Northern India during the second wave of the COVID-19 pandemic from May-June 2021. The study included 473 patients who tested positive for COVID-19. A high-resolution chest CT scan was performed within five days of hospitalization, and patient-related information was extracted retrospectively from medical records. Univariable and Multivariable analysis was done to study the predictors of poor outcome.

**Results:** A total of 473 patients were included in the study, with 75.5% being males. The mean total CT score was 29.89 ± 9.06. Fibrosis was present in 17.1% of patients, crazy paving in 3.6%, pneumomediastinum in 8.9%, and pneumothorax in 3.6%. Males had a significantly higher total score, while the patients who survived (30.00 ± 9.55 vs 35.00 v 6.21, p value - <.001), received Steroids at day 2 (28.04 ± 9.71 vs 31.66 ± 7.12, p value – 0.002) or Remdesivir had lower total scores (28.04 ± 9.71 vs 31.66 ± 7.12, p-value – 0.002). Total CT score (aHR 1.05, 95% CI 1.02 – 1.08, p – 0.001), pneumothorax (aHR 1.38, 95 % CI 0.67 – 2.87, p – 0.385), pneumomediastinum (aHR 1.20, 95% CI 0.71 - 2.03, p=0.298) and cardiovascular accident (CVA, aHR 4.75, 95% CI 0.84 – 26.72, p – 0.077) were associated with increased mortality, but the results were not significant after adjusting with other variables on multiple regression analysis.

**Conclusion:** This study identifies several radiological parameters, including fibrosis, crazy paving, pneumomediastinum, and pneumothorax, that are associated with poor prognosis in COVID-19. These findings highlight the role of CT thorax in COVID-19 illness and the importance of timely identification and interventions in severe and critical cases of COVID-19 to reduce mortality and morbidity.

## Introduction

Corona Virus Disease 2019 (COVID-19), caused by Severe Acute Respiratory Syndrome Coronavirus-2 (SARS-CoV-2) emerged as a global pandemic, threatening the health condition throughout the world with developing nations like India being the amongst the worst affected (1). With the accumulation of further evidence, certain clinical, laboratory and radiological factors were identified which could act as predictors of disease severity and predicting prognosis (2) (3) (4). The timely identification and interventions in severe and critical cases of COVID-19 are essential in reducing the mortality and morbidity of the disease (5).

The chest computed tomography (CT) scan plays a pivotal role in disease diagnosis, staging and in evaluation of treatment efficacy along with reverse transcriptase polymerase chain reaction (RT-PCR) which remains the mainstay for COVID-19 diagnosis (6). In a meta-analysis of 5744 patients the overall sensitivity, specificity positive predictive value, and negative predictive value of chest CT scan compared to RT-PCR were 87% (95% CI 85– 90%), 46% (95% CI 29–63%), 69% (95% CI 56–72%), and 89% (95% CI 82–96%), respectively (7). Another meta-analysis of 9907 confirmed COVID-19 patients showed that ground glass opacities, reticulations and air bronchograms were the most common chest CT findings (8). Certain Chest CT features also aid in staging and predicting severity of the disease (9) and bronchial wall thickening, GGOs, crazy paving and linear opacities were found to be associated with increased severity and mortality in one of the meta-analysis (10).

COVID-19 patients also develop certain complications like pneumomediastinum, pleural effusion, pneumothorax and pulmonary embolism over the ethics of illness which can significantly alter the ethics of hospital stay. Pneumomediastinum has been linked to poor clinical severity in one of the studies (11), however similar data on other parameters is lacking.

Thereby, this study is performed with the objective of finding chest CT parameters linked to prognosis in COVID-19 illness. It also aims to find demographic and lab parameters associated with radiological findings and poor clinical status.

## Methodology

This was a single centre hospital-based cross-sectional study conducted at a tertiary care centre in Northern India. Study included the patients who tested positive for either COVID-19 RT PCR/ COVID IgM antibody/ Rapid antigen for COVID 19 or the patient was clinically suspected to have COVID-19 infection. The study was conducted during second wave of Covid pandemic during the months of May-June 2021 and all the patients admitted in this time frame were included in the study. All the information related to the cases was extracted retrospectively from the medical records of patients.

All the patients underwent a high-resolution chest computed tomography (HRCT) scan within 5 days of hospitalization. Patient related information was extracted by trained physicians using standardized data extraction sheet. The data extracted included demographic variables like age, sex, addictions, comorbidities like hypertension, diabetes, coronary artery disease, chronic kidney disease, chronic liver disease, stroke/ TIA, obstructive airway disease. Data regarding HRCT thorax findings eg: extent of fibrosis, crazy paving pattern, pleural effusion, pneumo-mediastinum, pneumopericardium, pericardial/ pleural effusion, pulmonary embolism was collected from the radiology department and similarly symptom duration before admission, duration of hospital stay, maximum O2 requirement at admission, type of respiratory support (nasal prongs, face mask, non-rebreathing mask, high flow nasal cannula, non-invasive ventilation, invasive ventilation), lab parameters at admission eg: Hb, TLC, platelets, total bilirubin, SGOT/SGPT, urea/ creatinine, procalcitonin, D-Dimer, inflammatory markers like CRP, Ferritin, LDH were collected.

Correlation analysis was done using Spearman’s correlation coefficient. Univariable and multivariable logistic regression was done to assess association between various predictor variables and deaths due to COVID-19. A p<0.05 was considered as significant. Ethical approval for the study was obtained from the Ethics Committee of the All-India Institute of Medical Sciences, Rishikesh (CTRI/2020/08/027169).

## Results

A total of 473 patients were included in the study with 357 (75.5%) being males and 116 (24.5%) being females. The mean age of the study population was 52.04 ± 15.06 years. All the patients underwent a HRCT thorax and the mean CT scores were 7.96 ± 2.48 for the right lung, 7.92 ± 2.51 for the left lung and 29.89 ± 9.06 as the total score. Out of the fthis parameters assessed on CT thorax, 17.1% had fibrosis, 3.6% had crazy paving, 8.9% had pneumomediastinum and 3.6 % had pneumothorax. Amongst comorbidities, diabetes was the most common being present in 31.9% of all patients, followed by hypertension in 18% and coronary artery disease and COPD at 4.4% and 2.1% respectively. The mean O2 requirement was 2.63 ± 1.50 litres/min while the mean duration of hospital stay was 13.96 ± 9.51 days. 66.3% of the study population survived during the hospital stay (Table 1).

**Table 1.** Descriptive data of clinical characteristics of the cohort

| **Variables** | **N (%)** | **Mean (SD)** |
| --- | --- | --- |
| **Age (years)** | 403 | 52.04 (15.06) |
| **Right Score (of 20)** | 473 | 7.96 (2.48) |
| **Left score (of 20)** | 473 | 7.92 (2.51) |
| **Total score (of 40)** | 472 | 29.89 (9.06) |
| **Total days** | 337 | 11.47 (7.77) |
| **Haemoglobin (g/dl)** | 365 | 11.82 (2.43) |
| **Total leukocyte count (cells/cmm)** | 365 | 14.57 (14.42) |
| **Platelets (cells/nl)** | 364 | 210.85 (100.84) |
| **Bilirubin mg/dl)** | 77 | 3.88 (14.16) |
| **SGOT 9mg/dl)** | 77 | 93.16 (126.83) |
| **Urea (mg/dl)** | 101 | 70.83 (70.03) |
| **Creatinine (mg/dl)** | 98 | 2.28 (4.55) |
| **Procalcitonin (ng/ml)** | 78 | 6.49 (36.68) |
| **D-dimer (mg/L)** | 68 | 4.32 (6.30) |
| **O2 requirement (L/min)** | 324 | 2.63 (1.50) |
| **Duration of Hospital Stay (days)** | 389 | 13.96 (9.51) |
| **Sex (Male)** | 357 (75.5%) |  |
| **Fibrosis** | 81 (17.1%) |  |
| **Crazy Paving** | 17 (3.6%) |  |
| **Pleural effusion** | 62 (13.1%) |  |
| **Pneumomediastinum** | 42 (8.9%) |  |
| **Pneumothorax** | 17 (3.6%) |  |
| **Diabetes** | 151 (31.9%) |  |
| **Hypertension** | 85 (18.0%) |  |
| **Cerebrovascular Accident** | 3 (0.6%) |  |
| **Pulmonary Embolism** | 16 (3.4%) |  |
| **Outcome – Mortality** | 158 (33.7%) |  |

On bivariate analysis, males had a significantly higher total score (31.42 ± 7.59 vs 28.48 ± 9.98 in females, p value – 0.003). Total scores were higher in patients with comorbidities but the results were statistically insignificant. People who survived had a significantly lower score (30.00 ± 9.55 vs 35.00 v 6.21, p value - <.001). Patients who received steroids at day 2 had a significantly lower score (28.04 ± 9.71 vs 31.66 ± 7.12, p value – 0.002). The patients receiving Remdesivir had a lower O2 requirement (7.65 ± 2.64 vs 8.28 ± 2.07, p-value - 0.060) and lower Total score (28.04 ± 9.71 vs 31.66 ± 7.12, p-value – 0.002). Amongst all the CT parameters, pneumomediastinum and pneumothorax were associated with a higher mortality on bivariate analysis. Amongst all comorbidities, cardiovascular accidents (CVA) was associated with a higher mortality (Table 2 and Table 3).

**Table 2.** Bivariate analysis of CT scores and various clinical characteristics of the cohort

| Outcome | Male vs Female | Alive vs Death | Diabetic vs Non-diabetic | Steroids D2 vs No Steroids | Remdesivir vs No Remdesivir |
| --- | --- | --- | --- | --- | --- |
| **Right Score** | 3.11<br>(0.002) | -6.80<br>($<0.001$) | -1.766 (0.078) | 3.38 ($<0.001$) | |
| **Left score** | 2.03<br>(0.043) | -6.10<br>($<0.001$) | -0.989 (0.323) | 1.88 (0.060) | |
| **Total score** | 2.95<br>(0.003) | -7.24<br>($<0.001$) | -1.416 (0.157) | 3.11 (0.002) | 3.11 (0.002) |

N.B: The values in each cell show the T statistic and p-value for the comparison.

**Table 3.** Clinical characteristics of the cohort contributing mortality

| Factors | $\chi^2$ | df | p | $\epsilon^2$ |
| --- | --- | --- | --- | --- |
| **Sex** | 0.0161 | 1 | 0.899 | 4.04e-5 |
| **Fibrosis** | 0.3489 | 1 | 0.555 | 7.46e-4 |
| **Crazy paving** | 0.4418 | 1 | 0.506 | 9.44e-4 |
| **Pleural effusion** | 0.6630 | 1 | 0.416 | 0.00142 |
| **Pneumomediastinum** | 11.3354 | 1 | $<0.001$ | 0.02422 |
| **Pericardial effusion** | 3.0443 | 1 | 0.081 | 0.00651 |
| **Pneumothorax** | 7.5803 | 1 | 0.006 | 0.01620 |
| **Pulmonary embolism** | 0.7263 | 1 | 0.394 | 0.00156 |
| **Diabetes** | 3.1387 | 1 | 0.076 | 0.00671 |
| **Hypertension** | 0.4728 | 1 | 0.492 | 0.00101 |
| **Coronary Artery Disease** | 0.8255 | 1 | 0.364 | 0.00176 |
| **Cerebrovascular Accident** | 5.9304 | 1 | 0.015 | 0.01267 |

N.B: The Kruskal-Wallis test was used to compare the distribution of each factor between two groups. P-values less than 0.05 indicate a significant difference between the groups, while larger effect sizes ( $\epsilon^2$ ) indicate a stronger effect.

On correlation analysis, Total CT score had a weak positive correlation with the duration of hospital stay (spearman’s rho - 0.247, p<0.001) and a moderately positive correlation with O2 requirement (spearman’s rho - 0.416, p<0.001) (Fig 1).

**Fig 1.**
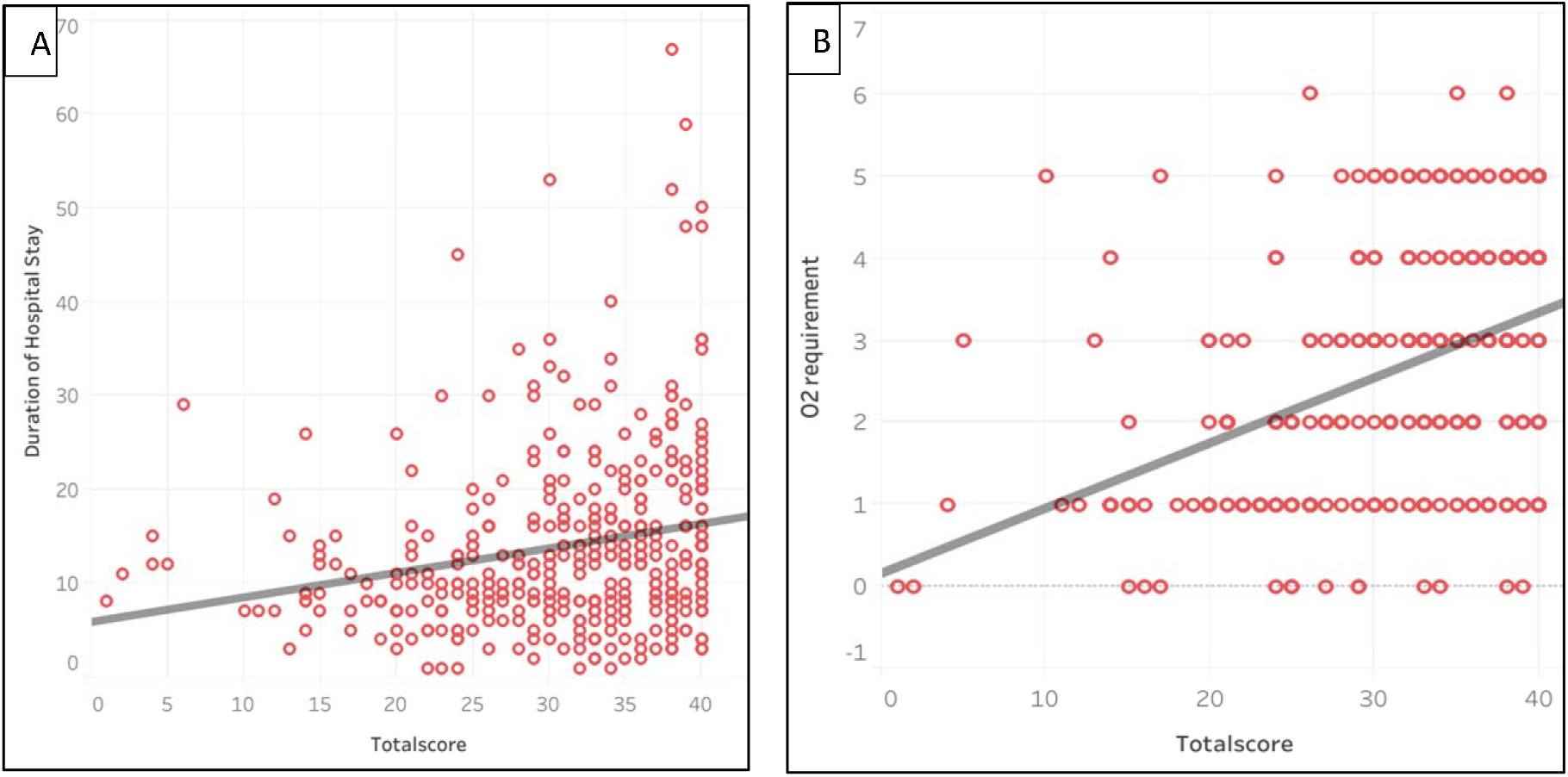
Correlation of CT score with duration of hospital stay (A) and O2 requirement (B). *Spearman’s correlation analysis was used to estimate the correlation equation.

On Multiple cox regression analysis, female sex (aHR 1.68, 95% CI 1.13 – 2.52, p – 0.011), CVA (aHR 4.75, 95% CI 0.84 – 26.72, p – 0.077), age (aHR 1.04, 95% CI 1.03 – 1.05, p <.001) and total CT score (aHR 1.05, 95% CI 1.02 – 1.08, p – 0.001) were associated with increased in hospital mortality. However, after adjusting for all the variables these were not associated with increased mortality (Fig 2).

**Fig 2.**
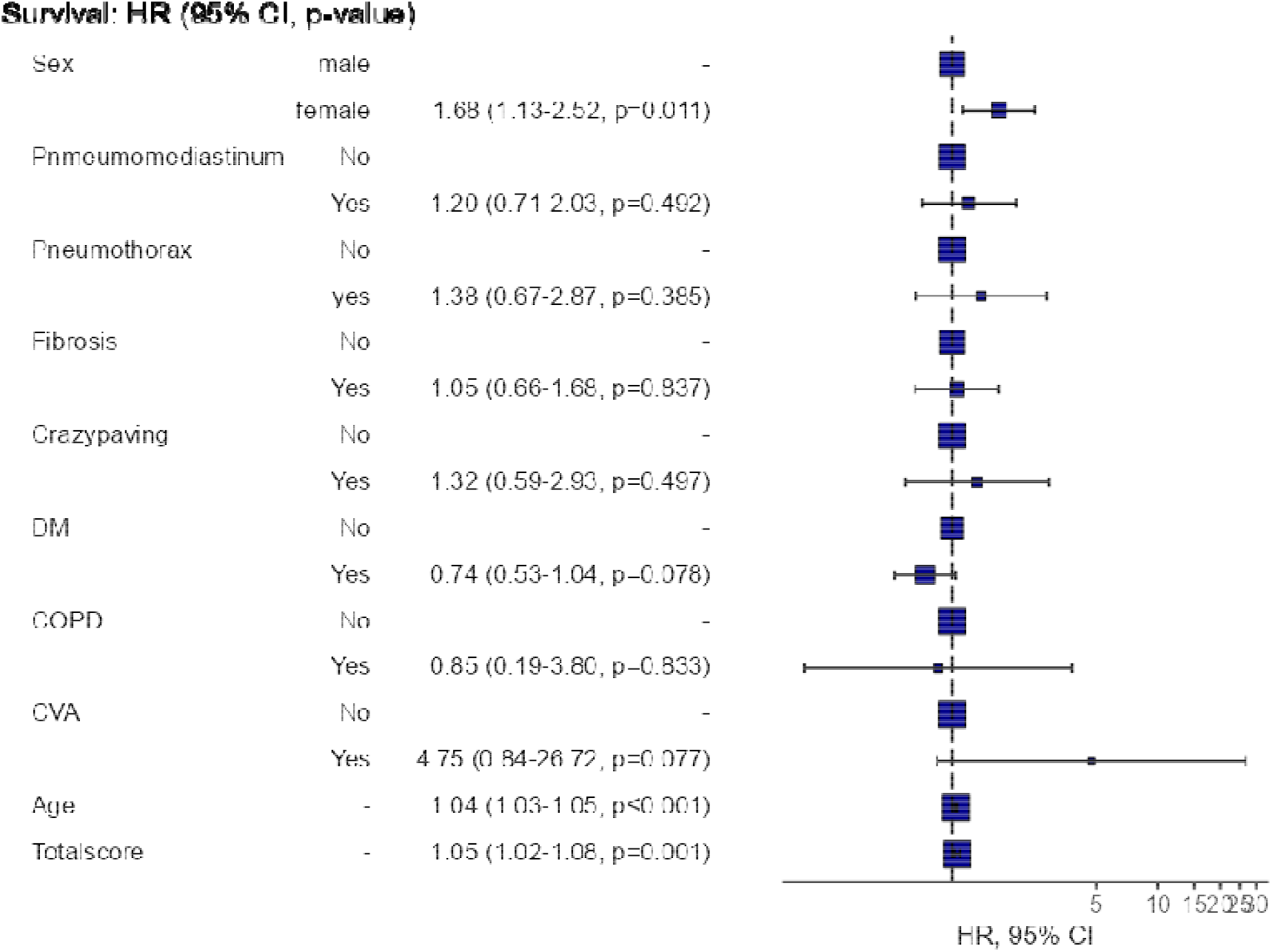
Results from the multivariable logistic regression model predicting in hospital mortality risk from COVID-19, presented as adjusted odds ratio with 95% confidence intervals. *Adjusted Odds Ratio, 95% Confidence Interval and p value of each as been mentioned.

## Discussion

The COVID-19 pandemic has led to a significant burden on healthcare systems worldwide, with high morbidity and mortality rates reported in severe cases. Various clinical and radiological parameters have been identified to predict the severity and prognosis of the disease. In this study, we found that fibrosis and crazy paving were common findings in COVID-19 patients and pneumomediastinum and pneumothorax were common pulmonary complications detected by CT thorax. We also found that CT severity scores were lower in survivors and the patients who received early steroids or Remdesivir. The duration of hospital stay and O2 requirement increased with increase in CT scores.

Recent studies have shown that COVID-19 primarily affects males, as seen in this study where 75.5% of the patients were male. This gender-based difference may be due to several factors, including differences in genetics, lifestyle, and immunity. For example, a study conducted in China found that the androgen receptor (AR) gene, which is responsible for regulating male hormones, could play a role in increasing the risk of COVID-19 infection and severity in men (12). Similarly, another study conducted in Italy found that male sex hormones may lead to a weaker immune response to COVID-19 infection, increasing the risk of severe disease (13).

In this study, the mean CT score for COVID-19 patients was 29.89 ± 9.06 which is similar to the findings of other studies, which have reported mean CT scores ranging from 18.5 to 32.8 (14). The most common CT findings in this study were fibrosis, crazy paving, pneumomediastinum, and pneumothorax and are consistent with previous studies, which have also reported similar findings. For instance, a study by Wang et al. reported that ground-glass opacities and consolidation were the most common CT findings in COVID-19 patients, and these were often accompanied by fibrosis, crazy paving, and pneumothorax (15). Similarly, another study by Xu et al. found that the most common CT findings in COVID-19 patients were ground-glass opacities and consolidation, and these were also accompanied by fibrosis and pneumothorax (16). Another study performed in the same institute showed correlation of increased CT scores with C-Reactive protein, d dimer and decreased lymphocytic counts (17). Therefore, this study confirms the common CT findings observed in previous studies and highlights the importance of using CT imaging for the diagnosis and monitoring of COVID-19 patients.

The presence of comorbidities has also been linked to increased severity and mortality in COVID-19 patients. This study found that diabetes was the most common comorbidity, present in 31.9% of all patients, followed by hypertension at 18%. Several studies have found that patients with pre-existing conditions such as diabetes, hypertension, cardiovascular diseases, and chronic respiratory diseases are at an increased risk of severe COVID-19 disease (16,18), but their association with worsened radiological scores or mortality was not found in this study and the literature on this aspect has been conflicting.

This study also found that patients who received steroids on day 2 or who received remdesivir had a significantly lower CT score, indicating less lung damage. This finding is consistent with the findings of a randomized controlled trial conducted by the World Health Organization (WHO) Solidarity trial, which found that systemic corticosteroids were associated with lower 28-day mortality in critically ill COVID-19 patients (19). Use of these drugs may have altered the CT scores by reducing inflammation.

Finally, this study found that several CT parameters, including pneumomediastinum and pneumothorax, were associated with increased mortality. These findings are consistent with the results of other studies that have identified pneumomediastinum and pneumothorax as indicators of severe disease and poor outcomes in COVID-19 patients (20,21). These CT parameters may be used as prognostic markers to identify high-risk patients who require close monitoring and aggressive treatment.

There were however a few limitations in this study. It’s a single centre study with a cross sectional design, which limits the ability to establish causality and results are not generalizable for a larger population. The study only assessed a limited number of laboratory parameters, which may not be sufficient to fully capture the complex pathophysiology of COVID-19.

## Conclusion

While some of the findings, such as the prevalence of comorbidities and their association with mortality, are consistent with previous studies, the specific findings related to CT scores and their correlation with age, O2 requirement, and duration of hospital stay are novel. Additionally, the study highlights the association of specific CT parameters like pneumomediastinum and pneumothorax with increased mortality, which adds to the existing literature on COVID-19. These findings could guide clinicians in the management and treatment of COVID-19 patients, especially in resource-limited settings. However, further studies are needed to validate this findings and investigate other potential risk factors for COVID-19 severity and mortality.

## Data Availability

All data produced in the present study are available upon reasonable request to the authors.

## Contributors

TC, AK, and AS contributed to the data collection, data analysis, and was involved in manuscript writing. PKP and SS gave the concept, interpreted analysis, critically reviewed the draft, and approved it for publication along with all authors.

## Data sharing

It will be made available to others as required upon requesting the corresponding author.

## Acknowledgment

None

